# Just4Us: Acceptability & Feasibility of a Woman-focused Pre-Exposure Prophylaxis (PrEP) Intervention: A Randomized Controlled Trial

**DOI:** 10.1101/2025.03.09.25323621

**Authors:** Hong-Van Tieu, Beryl Koblin, Vijay Nandi, Annet Davis, Ann Phan, Danielle Fiore, Anne M. Teitelman, the Just4Us Study Team

## Abstract

**Background:** Women comprise 19% of new HIV infections in the U.S. Many women, particularly women of color, are unaware of pre-exposure prophylaxis (PrEP) as an HIV prevention option. We conducted a pilot randomized controlled trial (RCT) to assess feasibility and acceptability of a theory-based, contextually-relevant behavioral intervention, Just4Us, to promote PrEP initiation and adherence among women in New York City and Philadelphia.

**Methods:** Eligibility criteria included: cisgender women, aged 18-55, not living with HIV, not currently taking PrEP, and meeting PrEP eligibility guidelines. Participants were randomized 3:1 to the Just4Us Education and Activities (E&A) arm or to the Information arm (Info); all were provided with PrEP information and a referral list. E&A arm participants received an individually-delivered session with a counselor-navigator, who provided information, motivation enhancement, skills-building, problem-solving, and referrals. Between baseline and 3-month follow-up, E&A arm participants received phone calls to support linkage to care and text-messages to promote adherence. Feasibility and acceptability were assessed.

**Results:** Eighty-three women were enrolled (61 intervention; 22 Info); 79% were Black, 26% Latina, exceeding diversity and enrollment targets. Attendance rate for the initial E&A and Info intervention session was 100%. Three-month retention rate was high at 90%. E&A arm participants reported feeling “very satisfied/satisfied with the following: overall session, 95%; discussion with counselor-navigator, 97%; tablet activities, 95%; text-messaging set-up, 93%.; and video, 90%. Among Info and E&A arms, 78% felt the session length was just right, and 95% stated that they would recommend/strongly recommend Just4Us to others.

**Conclusions:** The pilot RCT demonstrated feasibility and acceptability of the Just4Us E&A intervention, a promising intervention to increase uptake of PrEP among cisgender women. The team was able to recruit, implement the interventions with a high degree of fidelity, and retain the target number of PrEP-eligible socially disadvantaged women. Overall, participant feedback indicated they were generally very satisfied with their intervention.

## Introduction

In the United States (U.S.), 19% of new Human Immunodeficiency Virus (HIV) infections occur in cisgender women, with the most common mode of acquisition through heterosexual intercourse. Cisgender women of color are disproportionately affected. New infection rates in 2022 were nearly 8 times higher among Black women (15.5 per 100,000) and more than 3 times higher for Latina women (4.6), as compared to white women (1.9)[1].

Oral pre-exposure prophylaxis (PrEP) is an efficacious, self-administered, HIV prevention product. Among women who consistently take the medication, efficacy in preventing HIV acquisition ranges between 62% to 84%. Many women are unaware that PrEP is an available option for them to prevent HIV infection, and disparities exist by race/ethnicity in PrEP awareness and uptake that are disadvantageous to women of color[2]. The scale-up of PrEP is a prominent component of the Ending the HIV Epidemic Plan.[3]. Over a decade after oral PrEP was approved by the U.S. FDA[4], only 9.7% of eligible women who had indications for PrEP were prescribed PrEP in the U.S.[5].

There are currently few interventions in the U.S. to promote PrEP uptake among cisgender women, as the focus has primarily been on PrEP interventions among men who have sex with men and transgender women. Various approaches have been used to reach cisgender women in the U.S. Blackstock et al.[6] described a single arm pilot study conducted in New York City (NYC) examining an intervention called PrEP-UP, in which peer navigators provided support to cisgender and transgender women and transfeminine persons with PrEP education, counseling, navigation and linkage to care. A high proportion of the women (73%) expressed interest in taking PrEP, and 25% were assisted in scheduling PrEP appointments.

A study by O’Connell and Criniti assessed the feasibility of integrating PrEP counseling in a family planning clinic to improve knowledge and attitudes towards PrEP among women receiving family planning services in Philadelphia. Overall, knowledge and acceptability towards PrEP increased after receiving PrEP counseling. Participants who received more enhanced PrEP counseling guided by the Women’s PrEP Counseling Checklist (WPCC) experienced greater knowledge and acceptability of PrEP compared to women who received unguided counseling sessions[7]. Another single arm demonstration project study integrated PrEP with syringe services program in Philadelphia among women who inject drugs. The researchers found the women were interested and willing to start PrEP, and that streamlining it in an existing program can increase PrEP awareness and uptake and medical trust[8].

Researchers in Baltimore evaluated various PrEP promotion strategies in an obstetrics and gynecology clinic, serving primarily an African American population with access to care. They compared a nurse-led PrEP session with electronic clinic decision support tools (CSTs) on PrEP counseling and PrEP prescriptions. They found that PrEP counseling was higher in the group who received the nurse-led session (66.5%) compared to those in the active control arm with CSTs (12.3%) but PrEP prescriptions were similar between the two groups[9].

These studies indicate there are various ways to engage women in learning about PrEP and fostering informed PrEP decision-making. However, for women in the U.S., widely feasible, effective approaches to support use of PrEP are urgently needed. We conducted a pilot randomized controlled trial to evaluate the primary outcomes of feasibility and acceptability of a theory-based, contextually relevant, technology-enhanced behavioral intervention, Just4Us Education and Activities (E&A), to promote PrEP initiation and adherence among women in NYC and Philadelphia. The comparison group received a brief information session (Info). The secondary outcomes related to PrEP uptake behavior, intention and beliefs are reported elsewhere[10, 11]. The development of the Just4Us E&A intervention was informed by our multi-phase formative research, which involved 41 in-depth interviews and 160 surveys from cisgender women in both cities[12-15] as well as feedback from a community consulting group, as previously described[12].

## Methods

### Recruitment and Study Population

This study recruited and enrolled participants in Philadelphia and NYC from December 2018 to October 2019. Cisgender women were recruited using different approaches, including direct, in-person outreach at substance use programs and shelters, street outreach, placement of ads on social media and online websites, and referrals from enrolled women. Women were able to refer up to three women into the study, with reimbursement of $10 for each successful referral who enrolled in the study.

The eligibility criteria for the study were collected on a screening form based on identifying PrEP-eligible women and incorporated Centers for Disease Control (CDC) and New York State PrEP guidelines. Eligibility criteria included: cisgender women, aged 18-55 years, self-report being HIV-negative, not currently taking PrEP, and report having condomless vaginal or anal sex with a male partner or injected drugs in the past 3 months. In addition, the women had to report (a) either having or not knowing if they have a current male sex partner who injects drugs, who is HIV-positive, who has sex with men, or who was incarcerated in the past six months; or (b) reporting the following behavior in the past 6 months: shared injection equipment, used cocaine/crack cocaine, methamphetamines or Ecstasy at least once per week, been in a medication treatment program for opioid use (methadone, buprenorphine, suboxone), exchanged sex for money, drugs, and/or services, met the criteria for alcohol dependency (defined as a score of 2 or higher on the CAGE)[16], diagnosed with chlamydia, gonorrhea, syphilis or a new diagnosis of genital herpes, or had 3 or more male partners. Women were excluded from the study if they were currently pregnant, were currently taking PrEP, planned to relocate out of the area within the next 3 months, did not understand and read English at least a 5th grade level, did not have access to a mobile phone, or tested reactive on the rapid HIV test at the first study visit.

The study was reviewed and approved by the institutional review boards of the institutions involved. The study visits were conducted at the research sites in NYC and Philadelphia.

We included women of all racial/ethnic backgrounds, since any woman meeting the eligibility criteria should be provided with the opportunity to consider using PrEP. However, given the disparate impact of HIV on communities of color, we set a target of least 25-30% Hispanic/Latina and 65-70% Black/African American based on the racial/ethnic distribution of HIV infections among women in the two cities.

### Intervention and Comparison Group: Info and E&A Arms

Based on the Integrated Behavioral Model and the Theory of Vulnerable Populations, the Just4Us E&A intervention [12] included an in-person, individually-tailored, technology-enhanced session lasting 1 to 1.5 hours. The content was delivered by a Counselor-Navigator (C-N) at the baseline visit. The E&A arm curriculum had 12 mini-modules related to providing information about PrEP, motivation enhancement, skills-building, and problem-solving, and used different teaching modalities including video, tablet activities, and physical props. Between baseline and 3-month follow-up, C-Ns attempted at least 4 phone calls to support linkage to community-based PrEP care, using a person-centered problem solving approach to address any barriers. The same C-N who completed the intervention also completed the phone calls for a particular participant in the E&A arm to ensure continuity.

Just4Us participants utilized a simple automated text-messaging program based on the Twilio platform that sent women a self-customized confidential weekly query (created in the one of the last mini-modules of the intervention) on their mobile phones about whether or not they had started PrEP; if the woman replied “yes”, she began receiving her self-personalized daily reminder adherence messages, followed one hour later by a confidential prompt to reply “yes” or “no” (indicating if she had taken her pill for the day).More details about the E&A and Info intervention arms have been previously described[12].

The comparison group (henceforth “Info arm”) participants met individually with a C-N in an in-person the baseline visit. During the 5-10 minute session, the C-N provided a brief description of the handouts in the packet. Participants in both arms received a packet of handouts on PrEP facts, cost, and PrEP providers, along with other materials about maintaining healthy lifestyles, nutrition, smoking cessation, and breast cancer screening. All sessions in both arms were audio-recorded.

### Counselor-Navigator (C-N) Training

The C-Ns (3 in NYC, 3 in Philadelphia) were provided with a 6-hour, in-person group training that included (a) an overview of the Just4Us E&A arm, including theoretical approach and formative research; (b) a description of facilitation strategies, such as motivational interviewing techniques and perspectives such as harm reduction and empowerment; (c) a detailed explanation of each E&A intervention module with time to role-play activities; (d) overview of the content of the Info arm. Facilitator guides for both arms were distributed to all C-Ns to use as a reference and included written descriptions of training content, activities, and handouts.

### Baseline, Post-intervention, and Follow-up Study Visits

After providing written informed consent at the baseline visit, all participants viewed a brief informational video about PrEP for women, completed a computer assisted self-interview (CASI) baseline questionnaire, and received HIV counseling and a rapid HIV test from a trained HIV counselor/data collector. They were then randomized in a 3:1 ratio into the E&A arm or Info arm, stratified by site; the randomization process is described elsewhere[17].

Participants then met with a C-N who delivered the material for the E&A arm (consisting of information, counseling, and navigation) or Info arm. After the intervention session, participants met again with the data collector to complete the post-intervention CASI questionnaire focused on assessing social-cognitive covariates of PrEP uptake as well as eliciting feedback about the intervention session. At the 3-month follow-up visit, participants completed a CASI questionnaire and a brief exit interview. Participants were reimbursed $50 for their time and travel for the baseline visit and $50 for the 3-month follow-up visit.

## Data Collection and Measures

### Eligibility Screening and Enrollment Data

During the study enrollment period, the database manager (DF) monitored all screening and eligibility forms at both sites and summarized data onto an excel spreadsheet. This data was reviewed and monitored at our weekly team meetings. Tracked information included eligibility criteria responses and race/ethnicity for all those who were screened, status (non-eligible/eligible/enrolled/consented) and relevant dates (screening and enrollment).

### Participant Data

#### Baseline Survey

The participants completed a baseline CASI survey, with questions pertaining to sociodemographic information including age, race/ethnicity, educational level, income, employment, and financial instability. HIV-related behaviors including injection drug use, condom use with vaginal and anal sex partners, partner concurrency and transactional sex; perceived HIV risk; drug use in the past 3 months; and PrEP awareness. PrEP knowledge and social cognitive variables pertaining to PrEP uptake were also assessed. The survey also collected information about housing status, intimate partner violence, self-help skills, phone and internet access, transportation access, and access to social and legal services and housing assistance.

#### Post-Intervention Survey

The post-intervention survey, which was completed at the end of the baseline visit, included general questions about satisfaction with their intervention arm: (1) How helpful was your session with the C-N? (4-point Likert scale, not at all helpful to very helpful), (2) How enjoyable was your session with the C-N? (4-point Likert scale, not at all enjoyable to very enjoyable), (3) How clear was the information in your session with the C-N? (4-point Likert scale, not at all clear to very clear), (4) What do you think about the duration of your session with the C-N (not including the survey on the computer)? (3-point Likert scale, too short, just right, too long), (5) In general, how much did you learn from the session with the C-N? (4-point Likert scale, learned nothing to learned a lot), (6) Overall, how satisfied are you with your session with the C-N? (4-point Likert scale, from not at all satisfied to very satisfied), and (7) Would you recommend Just4Us to others? (4-point Likert scale, would not recommend to would strongly recommend).

In addition, two open-ended questions were asked: “What did you like about your session with your C-N? Please explain” and ‘What did you not like about your session with your C-N? Please explain”. More specific questions about various intervention components were asked of those randomized to the E&A arm, including (1) How satisfied are you with the video at the beginning of the session? (4-point Likert scale from not at all satisfied to very satisfied), (2) How much did you like the activities on the tablet when you were meeting with the C-N? (4-point Likert scale, disliked very much to liked very much), (3) How satisfied are you with the discussion with the C-N?, (4) How satisfied are you with setting up the text messaging with the C-N?, (5) How satisfied are you with the video at the beginning of the session?, (6) How much did you like the activities on the tablet when you were meeting with the C-N? (4-point Likert scale, disliked very much to liked very much), (7) How satisfied are you with the discussion with the C-N?, and (8) How satisfied are you with setting up the text messaging with the C-N?. Responses were given in 4-point Likert scale, from not at all engaged to very engaged.

#### Three-Month Follow-up Survey

At their last study visit, the participants completed a survey similar to the baseline survey with the addition of a few questions related to the interventions (e.g., those in the E&A arm were asked if they had received the follow-up text messages and to rate their satisfaction with them). In addition, the data collector conducted the brief exit interview by asking several open-ended questions and typing in participants’ responses on an online form. The opening question asked: “Tell me about your experiences in the past few months as you were thinking about starting PrEP?” The data collector asked follow-up probes, such as, “Can tell me more about that?” In the first few exit interviews, the women referenced talking to others about PrEP, thus in subsequent interviews, the study team added follow-up questions (e.g., “Did you talk to anyone about PrEP?”) to gain more insights.

### Counselor-Navigator Documentation

The C-Ns completed an online Log Sheet in Qualtrics after each intervention session, noting what intervention (E&A or Info) content was completed.

For the 4 follow-up phone calls, the C-N used a discussion guide to document the phone calls; First the C-N assessed the participants progress in acquiring PrEP. Specific questions (with ‘yes’/‘no’ responses) were asked based on their place on the PrEP cascade, e.g., (a) *before your visit*: (1) Did you find a provider?, (2) Did you find out about how to pay for PrEP? (3) Did you schedule your appointment?; (b) *during your visit:* (1) Were you able to go to your appointment?, (2) Did you ask your provider to write you a prescription for PrEP?, (3) Were you able to get lab work drawn to know if it’s safe and effective for you to start PrEP?; (c) *after your appointment:* (1) Did you need to call your provider to check if your lab results are ready?, (2) Did you return to your provider to discuss the lab results?, (3) If you were cleared to start on PrEP, did you ask for a prescription at this visit?, (4) Were you able to get your prescription filled at a pharmacy?, (5) Were you able to pick up the prescription at the pharmacy?, (6) Were you able to start taking a pill once a day?, and (7) In addition, the C-N asked participants open-ended questions about any challenges, specific barriers, and developed an action plan for each specified barrier.

### Assessments of Counselor-Navigators

After the group training session, the C-Ns conducted a mock C-N session with a co-worker on video followed by a self-evaluation. Two members of the research team (AMT & BAK) independently evaluated each video using a standardized assessment form. To proceed to working with study participants, the C-Ns had to achieve a mastery of 4–5 (very good–excellent) on all components, repeating the mock C-N session on video as needed. While working with study participants, C-Ns received ongoing supervision, with case review and problem solving of issues that arose during the interviews (i.e., what worked well, areas of improvement), provided during weekly team meetings.

Fidelity and quality of intervention sessions were assessed using the C-N Intervention Evaluation Questionnaire, a standardized, 5-point Likert rating form (poor to excellent) completed by two study team investigators (AMT & BAK), who listened to the audio recordings of selected intervention sessions of both arms and across all C-Ns. The following categories were assessed: presentation skills (8 items), knowledge and communication skills (13 items for E&A arm; 11 items for Info arm), and adherence to module objective(s) depending on the arm (1 item for the Info arm; 12 items pertaining to each module for the E&A arm). The C-Ns were given their completed fidelity and quality rating forms as part of ongoing supervision and feedback during the study.

### Data Analysis

We determined feasibility and acceptability outcomes as follows.

**Feasibility** was assessed by the following measures: (1) number of women eligible relative to number screened, (2) number enrolled relative to number eligible, (3) consent rate (defined as the number of eligible women successfully consented for intervention relative to those who were eligible but who did not consent), and (4) number enrolled relative to the target sample size of 80. Feasibility benchmarks included at least 40% eligible:screen ratio, at least 75% enroll:eligible ratio, 100% consent rate, and reaching or surpassing target enrollment of 80 women. We documented the enrollment period in months to recruit and enroll the target sample size of 80 women. We also assessed feasibility of enrolling a target of least 25-30% Hispanic/Latina and 65-70% Black/African American women in the study, by screening for race/ethnicity in study enrollment

We qualitatively evaluated the exclusion reasons for women were not eligible and inclusion categories of the women who were eligible, and compared baseline demographics and PrEP-relevant behaviors to determine any differences between eligible women who enrolled and who did not enroll into the study.

In addition, we evaluated the feasibility of intervention training. Counselor-navigator training evaluations were used to indicate mastery, with a benchmark of 100%.We assessed intervention fidelity and quality, with benchmark ≥ 90%, as defined by the percent of modules and required behaviors checked on C-N log sheets as completed appropriately and quality rating of 4 (good) or 5 (excellent) on the C-N Intervention Evaluation Questionnaire

**Acceptability** was assessed by: E&A intervention completion rate, 3-month visit retention rate (defined as the number of women who completed the 3-month visit as a proportion of the total number of participants enrolled into the study), and satisfaction level based on survey responses on the post-intervention CASI survey completed at the end of the baseline visit.

Binary measures were created by grouping response levels for outcomes and behaviors with multiple response levels. Binary variables were derived as follows: for the 4-point Likert scale questions, the two favorable responses were combined and the two unfavorable responses were combined; for the length of session question “just right” was designated as the favorable response and “too short"/"too long” were combined to be the unfavorable response.

Acceptability benchmarks included attendance rate for the initial intervention of 100% and intervention completion of at least 90% (defined as the percent of participants who attended baseline session and completed all the modules). We assessed completion of the phone call components (defined as uptake of PrEP before 4 calls or completing 4 calls within 8 weeks). In addition, we had a benchmark retention rate of at least 90% ( defined as percent of participants who completed 3-month follow-up assessment relative to those who were lost to follow-up). Benchmarks for satisfaction levels of the E&A and Info arms included at least 80% of participants reporting moderate or higher levels of satisfaction and acceptability on a 4-point Likert scale on the Brief Satisfaction Questionnaire.

Descriptive statistics were used to summarize the participants’ sociodemographic variables, risk behaviors, perceived risk, and PrEP-relevant behaviors. Exact binomial tests were used to generate proportions and 95% confidence intervals (CIs). All statistical analyses were conducted using R version 4.0.4, and figures were created in Microsoft Office Excel (365).

Content analysis was used to assess the data from the open-ended participant and C-N survey responses and the brief exit survey[18]. Initially, the data were reviewed, and broad topics were identified and coded by two study team members. The code book was shared at larger team meetings for input and consensus was reached on the codes though discussion. Two study team members identified nuances within codes using the constant comparison approach, and created relevant descriptive subcategories, discussing any discrepancies until consensus was reached.

## Results

### Sample Characteristics

At baseline (Table 1), the mean age of the 80 women included in the analysis (who enrolled, were allocated to intervention or control, and completed post-intervention survey) was 37 years, 63 (78.8%) identified as Black/African-American, and 21 (26.3%) as women of Hispanic/Latina ethnicity. This group of PrEP-eligible women faced significant challenges on multiple levels. Most (82.5%) reported that they sometimes or very often did not have enough money for basic necessities (such as rent, food, utilities) in the past 3 months.

**Table 1:** Participant Baseline Sociodemographic Characteristics and Risk Behaviors.

| **Characteristic** | **Total**<br>**(n=80<sup>a</sup>)** | **Intervention**<br>**(n=58<sup>a</sup>)**<br>**(E&A)** | **Control**<br>**(n=22)**<br>**(Information only)** |
| --- | --- | --- | --- |
|  | **Mean (SD)** | **Mean (SD)** | **Mean (SD)** |
| Age (years) | 37 (12) | 37 (12) | 39 (13) |
|  | **N (%)** | **N (%)** | **N (%)** |
| Race |  |  |  |
| African American | 63/80 (78.8) | 46/58 (79.3) | 17/22 (77.3) |
| White/Other | 17/80 (21.2) | 12/58 (20.7) | 5/22 (22.7) |
| Hispanic/Latina ethnicity | 21/80 (26.3) | 16/58 (27.6) | 5/22 (22.7) |
| Annual Income |  |  |  |
| Less than \$12,000 | 56/79 (70.9) | 41/57 (71.9) | 15/22 (68.2) |
| Past 3 months had experienced financial insecurity | 66/80 (82.5) | 47/58 (81.0) | 19/22 (86.4) |
| Education |  |  |  |
| Less than high school diploma | 22/80 (27.5) | 17/58 (29.3) | 5/22 (22.7) |
| A high school diploma or GED | 27/80 (33.8) | 18/58 (31.0) | 9/22 (40.9) |
| Some college/trade school, or more | 31/80 (38.7) | 23/58 (39.7) | 8/22 (36.4) |
| Employed | 20/80 (25.0) | 16/58 (27.6) | 4/22 (18.2) |
| Current perceived HIV risk | 41/80 (51.2) | 29/58 (50.0) | 12/22 (54.5) |
| PrEP relevant behaviors in the last 3 months: |  |  |  |
| Any drug use | 41/80 (49.2) | 29/58 (47.5) | 12/22 (54.5) |
| 3 or more alcohol drinks per day | 30/80 (37.5) | 25/58 (43.1) | 5/22 (22.7) |
| Had condomless sex (vaginal or anal) | 52/77 (62.5) | 33/55 (54.1) | 19/22 (86.4) |
| Had multiple partners (more than 1 partner) | 59/80 (71.1) | 42/58 (62.9) | 17/22 (77.3) |
| Engaged in sex for money, drugs, goods, or other services | 37/78 (47.4) | 29/56 (51.8) | 8/22 (36.4) |
| Intention to start PrEP in the next 3 months | 63/80 (78.8) | 46/58 (79.3) | 17/22 (77.3) |
| Heard of PrEP | 38/80 (47.5) | 25/58 (43.1) | 13/22 (59.1) |

<sup>a</sup> Baseline surveys were missing from 2 women in the E&A arm. Total number of women who completed the baseline surveys was 80 women: 58 in the E&A arm and 22 in the Info arm.

Alcohol and other drug use was prevalent, with 37.5% of the women reporting having 3 or more drinks per day and 49.2% reporting any drug use. Sexual behaviors that increased the likelihood of HIV exposure were also common, as 71.1% had more than one partner in the past 3 months and 62.5% reported condomless vaginal or anal sex in the past 3 months. Over two-thirds (68.0%) reported using drugs or alcohol to get high or drunk before having sex in the prior 3 months, and 47.4% engaged in sex for money, drugs, goods or other services. Slightly over half (51.2%) believed they were at risk of acquiring HIV in the prior 3 months. Slightly more than half the women (42/80; 52.5%) had not heard of PrEP prior to participating in this study. After PrEP information was provided, more than three-quarters (78.8%) indicated that they intended to use PrEP in the next 3 months. When asked about their preference for PrEP modality, 56% preferred a daily pill, 29% injectable, and 15% did not know.

In addition, 90% (72/80) of the women in the two arms reported having access to a smartphone, with only 1 woman reporting sharing her device with another person. Furthermore, 72.5% (58/80) reported having access to unlimited data plan, while 46.3% (37/80) indicated that, in the past year, their phone service had been disconnected because of inability to pay the bill or because the phone was lost or stolen.

## Main Outcomes

### Feasibility: Recruitment, Enrollment and Retention

A total of 255 women were recruited and assessed for eligibility for the study (Figure 1: Consort Flow Diagram). Of 255 screened, 172 women were excluded, with 151 women who were found to be ineligible and 21 who were unable to schedule a study visit or did not show to the appointment. Among the 151 ineligible participants, 89 did not report having condomless vaginal or anal sex with a male partner in the past 3 months or injecting drugs in the past 3 months; 98 did not meet additional risk criteria; 16 did not have a cell phone; 14 were excluded due to race/ethnicity aims (identified as White, non-Hispanic/Latina); 9 did not meet age criteria (18-55 years); 6 were pregnant; 5 did not understand English; 4 were already on PrEP; 3 did not identify as women; and 3 reported being HIV positive.

**Figure 1:**
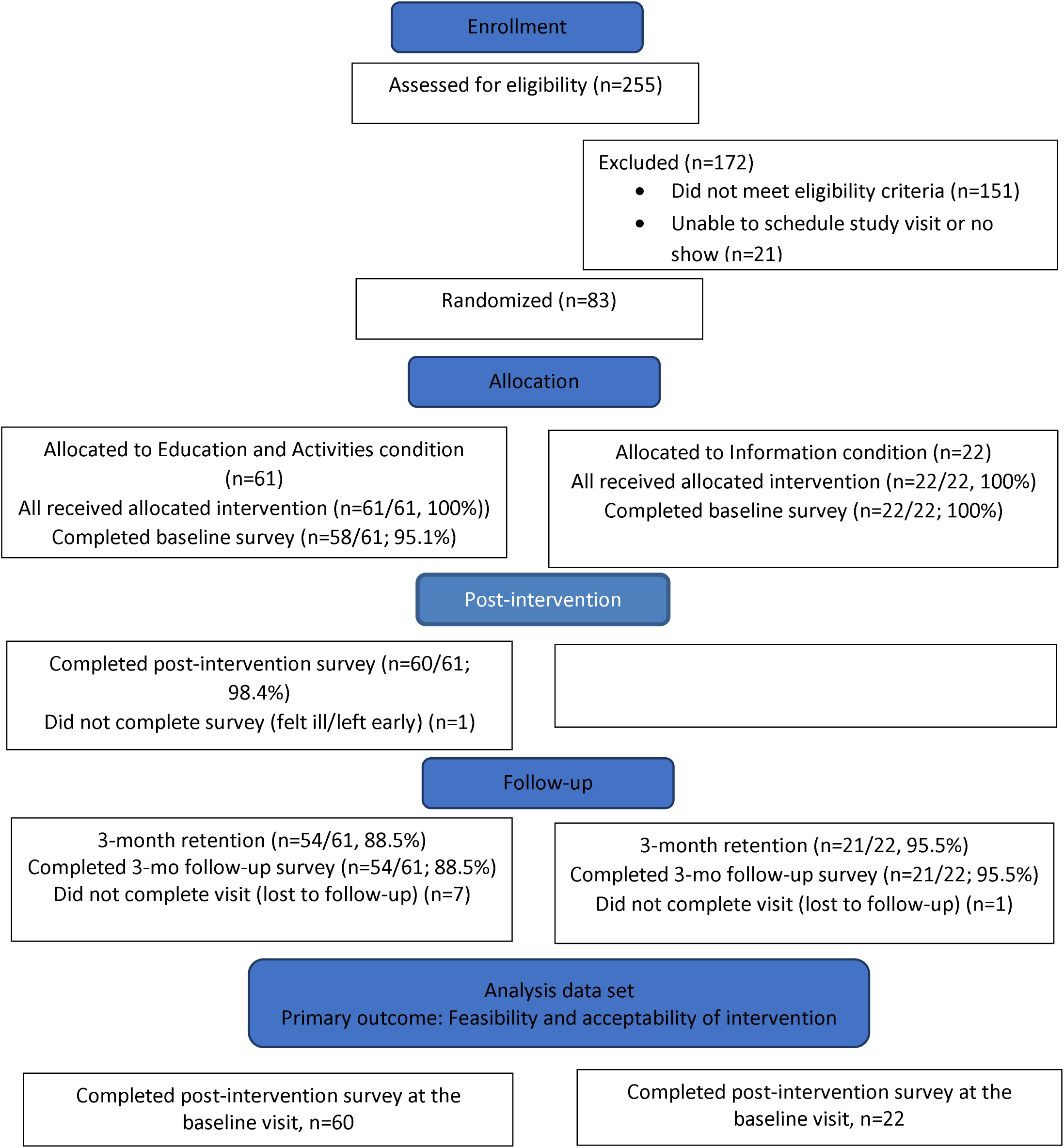
Consort Flow Diagram. Consort flow diagram for randomized control trial of Just4Us intervention among E&A and Info arms

The remaining 104 women (41%) were eligible, and 83 (80%) were enrolled in Philadelphia or NYC (61 in the E&A group and 22 in the Info group), exceeding target enrollment of 80 to account for lost baseline surveys from 3 participants in the E&A arm (Figure 1). Among the 83 women enrolled, the most common recruitment sources were referrals from enrolled participants (47.0%), direct outreach at transitional housing for post-prison release, substance recovery programs, and shelters (28.9%), and online ad postings (e.g., Craigslist) (9.6%).

The feasibility benchmarks of at least 40% eligible:screen ratio, at least 75% enroll:eligible ratio, and reaching or surpassing the target enrollment of 80 women were met. All (100%) of the eligible & enrolled women successfully consented for the study, meeting the benchmark. The 83 women were recruited and enrolled over an 8-month period from Nov 2018 to June 2019, with 40 in Philadelphia and 43 in NYC. Participant accrual was slow in the first two months, with 4 enrolled, but then increased to approximately 13 participants per month over the subsequent 6 months.

Of the 61 women allocated to the E&A arm, all received the intervention; however, the completed baseline surveys from 3 women were missing, and one woman felt ill and did not complete the post-intervention survey (Figure 1). Of the 22 women randomized to the Info arm, all received Info, and all completed the baseline and post-intervention surveys. The analysis data set for most of the feasibility benchmarks, therefore, included a total of 82 women: 60 in the E&A arm who completed the E&A intervention and post-intervention survey and 22 in the comparison group who completed Info intervention and post-intervention survey. The analysis data set for the 3-month retention rate and text-message delivery analysis was n=75 (54 E&A; 21 Info).

We exceeded our enrollment targets by race/ethnicity, with 26.3% of the women identifying as Hispanic/Latina and 78.8% identifying as Black/African American. We compared key baseline demographics between all 104 women who were eligible (including those who enrolled and did not enroll) and 83 women enrolled in the study. Among the 104 eligible women, the mean age was 36.9 years (SD 12.0); 69.2% were Black and 25.0% were Hispanic. Among the 83 women who were eligible and enrolled, the mean age was 37.7 years (SD 12.1); 78.8% were Black and 26.3% were Hispanic.

As reported previously[17], the 3-month visit was completed by 75 participants, resulting in a high overall retention rate of 90.4% at 3-month follow-up, meeting our benchmark. Retention was 88.5% in the E&A arm and 95.5% in the Info arm (Fisher’s exact test, two-sided p-value p=0.675). At 3-month follow-up 54/61 E&A participants and 21/22 Info participants competed the survey.

### Acceptability

#### Intervention attendance rate and module completion

The baseline survey data were collected on 58/61 Just4Us E&A participants (95%) and all 22 Info session participants, resulting in an overall 96.4% baseline survey completion rate across the two arms

Attendance rate for the initial E&A and Info interventions was 100%, meeting an acceptability benchmark. Among the 60 women randomized to the E&A intervention and completed the post-intervention survey, completion of the 12-module intervention was nearly 100%: 10 modules had a 100% completion rate, 4 participants did not complete the text messaging module, which included an overview of text messaging for PrEP and enrollment in a text messaging program (overall 56/60 or 93.3% completion), and one participant did not complete the final module involving reinforcement of the PrEP plan (overall 59/60 or 98.3% completion). This met our module completion benchmark of at least 90%. As previously reported[17], an evaluation of the audio recordings from the 21 sessions (14 E&A intervention sessions, 17 Info arm sessions) showed that the mean intervention ratings were 4.8/5 (SD=0.29) for quality and 4.54 (SD=0.52) for fidelity. In comparison, the Info arms sessions had mean ratings of 4.98 (SD = 0.04) for quality and 5.00 (SD = 0.00) for fidelity.

#### Text messages

At baseline, 4 participants did not complete the text messaging module or indicated they did not want to participate in the text messaging program; however, all 4 completed the 3-month follow-up survey and are included in this summary.

Based on the survey, 78% (42/54) of the E&A arm and who completed the 3-month follow-up visit reported receiving the weekly text messages; of those, 85% expressed that they were satisfied/very satisfied with them. Of the 6 women who started PrEP in the E&A arm, only half indicated they received the daily reminders.

#### Post-intervention telephone call follow-ups

As noted above, the C-Ns completed an online form after each call/attempt to call (for E&A only). A total of 81 calls were documented by staff at both sites to 39 participants, with a range of 1 to 14 calls per participant (median 2 calls).

#### Reasons for unsuccessful calls

C-N notes about the reasons for unsuccessful calls were available for 30 of the 39 participants with documented call attempts. These included unanswered calls or calls going to voicemail, which even happened when the C-N had previously spoken to the participant who had agreed to a specific time to call. The C-Ns would also sometimes use text messages to reach participants to reschedule a follow-up call.

#### Satisfaction levels with intervention components and control arm

Figures 2 and 3 describe the responses in the post-intervention survey in the E&A arm and both arms, respectively.

**Figure 2:**
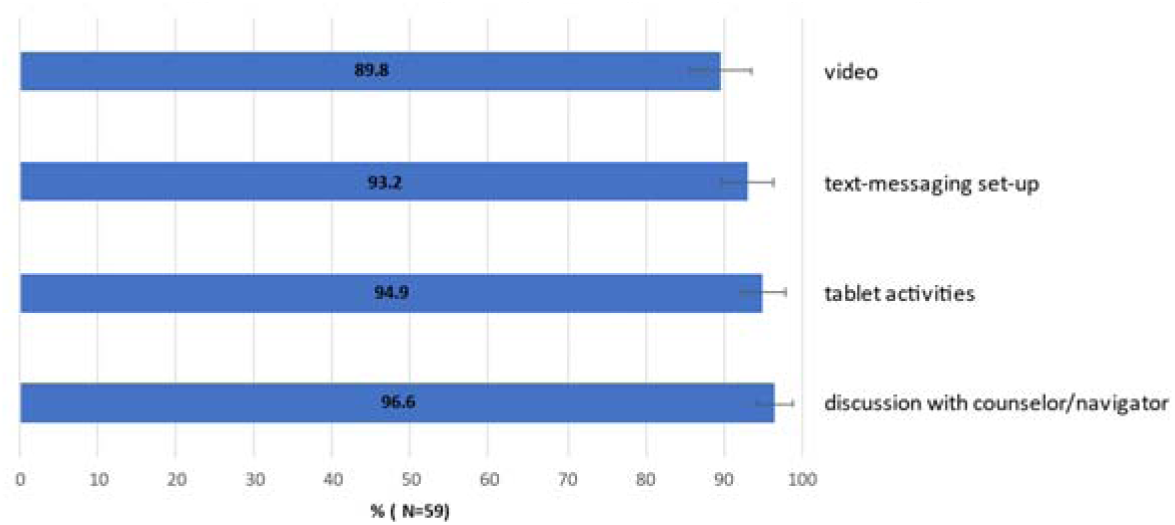
Intervention participants reporting “very satisfied/satisfied” levels. Percentage of E&A arm participants reporting “very satisfied/satisfied” levels for different components of intervention

**Figure 3:**
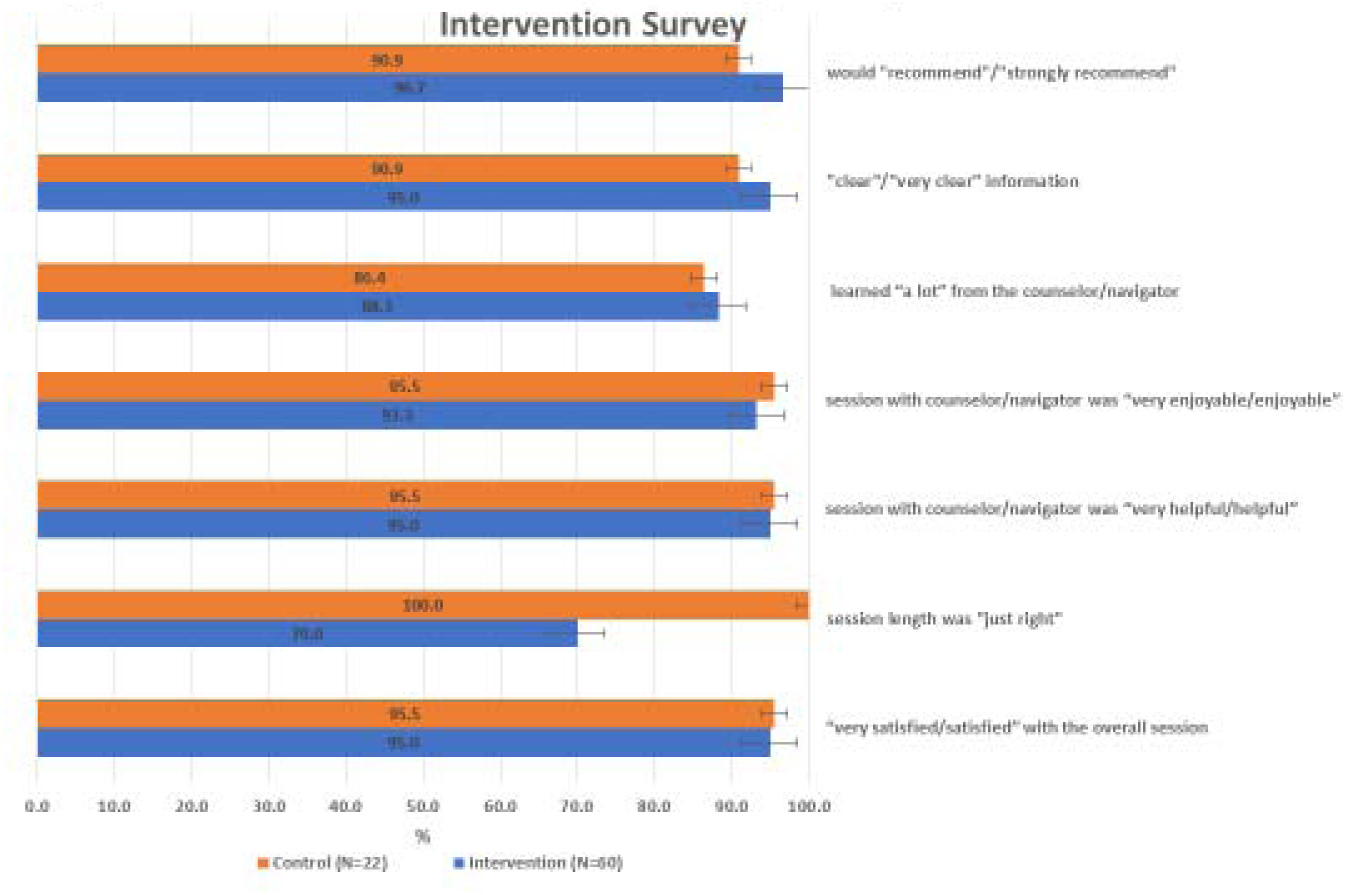
Intervention and Control Arm Participant Responses on Post-Intervention Survey. Comparison of participant responses to post-intervention survey for E&A and Info arm participants. The orange represents the Info arm, while the blue represents the E&A arm.

Participants in the E&A arm reported the following “very satisfied/satisfied” levels: overall session, 94.9%; discussion with C-N, 96.6%; tablet activities, 94.9%; text-messaging set-up, 93.2%; video, 89.8% (Figures 2 and 3). A majority of participants in both the E&A and Info arms indicated they learned a lot from the C-N (86.4% Info vs. 88.3% E&A); they also found the session to be very enjoyable/enjoyable (95.5% Info vs. 93.3% E&A) and very helpful/helpful (95.5% Info vs. 95.0% E&A). Regarding session length, 78.0% of participants felt it was just right, with statistically significant differences between the arms: 100% of Info arm participants rated the session length as “just right,” while this percentage dropped to 70.0% among E&A participants. Overall, nearly all (95.1%) of E&A and Info participants reported being satisfied or very satisfied with the session (95.5% Info vs. 95% E&A). Similarly, almost all respondents (95%) stated that they would recommend or strongly recommend Just4Us to others, with a slightly higher percentage among E&A participants (96.7%) compared to Info arm participants (90.9%).

### Feedback from the women based on answers to the two open-ended post-intervention survey questions (n=82)

Participants’ responses regarding what they liked about their sessions with the C-N showed some similarities across both groups. The most frequent comments highlighted qualities of the C-N, such as being friendly, nice, knowledgeable, or considerate. Many described their interactions with the C-N as comfortable, with statements such as, “[The C-N was] friendly and made me comfortable about speaking about my personal life” (Info), or “I felt comfortable in discussing personal information” (E&A). Both groups emphasized that the information was presented clearly. For example, a participant in the Info arm noted, “She was very personable, and clear on words. She was also considerate when asking personal questions and had really great manners. She was really nice!”

Notably, participants in the E&A arm mentioned more specific aspects they appreciated, such as the detailed explanations: “I like how she explained it to step by step” and “I appreciated the time she took to explain PrEP and the benefits of taking it”. Some participants highlighted the personalized nature of the sessions, stating that “[the C-N] taught me a lot about PrEP and made PrEP useful for me” or “I liked that she took her time to explain everything to me, she made sure I understood everything” or “The way she explained my decisions/choices about safer sex, and including the use of PrEP medication.” In addition, participants in the E&A arm noted the activities were fun, interactive and beneficial. Comments included, “She was funny but yet still very educational. This is how I learn” and “I liked the role play when telling my healthcare provider, ‘I’m interested in starting PrEP.”

When asked what they did not like about their session with the C-N the most common response was “nothing”. A couple of participants in the Info arm wanted more detailed information about PrEP, “I would’ve liked a little more information on prep.” Twelve out of sixty-one participants (20%) in the E&A arm (J4U) indicated the session was too long.

### Exit interview responses after completion of the 3-month follow-up survey (n=75)

In the exit interviews conducted at the end of the 3-month follow-up, some participants in both arms provided general feedback about the intervention session (21 participants in the E&A arm and 9 in the Info arm). Consistent with the post-intervention survey responses described above, participants generally expressed that they enjoyed the intervention and valued the information shared. A few noted the need for PrEP in their community, emphasizing that the intervention was a good way to educate the public about PrEP. Unexpectedly, some women mentioned that after participating in the intervention, they “often share my knowledge [about PrEP] with others” This prompted us to probe for any discussions they may have had with others about PrEP. Twenty-seven of the participants in the E&A arm reported that they talked to their family, friends, and partners about PrEP, including some individuals who used substances. Overall, most reactions from those they spoke to were positive towards PrEP, with comments including “better to be safe than sorry” and “it is a good idea”. Among those supportive of PrEP, some mentioned that it was their first time learning about it, while others expressed concerns about potential side effects.

In sharing PrEP information with friends and others, the participants framed PrEP as a way to reduce the risk of acquiring HIV, especially in high-burden communities. One participant noted, “It will prevent you from HIV and in our community, there has been a lot [of HIV]”, while another added, “People who use drugs might have unprotected sex”. One participant stated, “I have been telling some friends and my daughter because of drugs and all that’s going on…I just wish PrEP had been around sooner to help others”.

Seven E&A participants reported negative reactions when discussing PrEP with others. These primarily were related to perceptions that PrEP was intended for individuals who are promiscuous, being too old, potential side effects and stigma. One participant shared a friend’s comment: “If you talk too much about it [HIV], you may get it.”

Among the 9 participants in the Info group who discussed PrEP with others, one reported a positive reaction, three negative reactions (e.g., concern for side effects), while the others were neutral (e.g., needed more information).

## Discussion

The HIV epidemic continues to disproportionately affect cisgender women of color living in low-income urban areas, including Philadelphia and NYC. While PrEP is highly effective in preventing HIV infection when taken consistently, awareness and uptake among women at risk of exposure to HIV remain very low. Currently, there are few interventions in the US aimed at promoting PrEP uptake among eligible cisgender women. Therefore, effective approaches to support PrEP uptake among women in the U.S. are urgently needed.

In this pilot randomized controlled trial involving a theory-based, contextually relevant, technology-enhanced behavioral intervention, Just4Us, to promote PrEP initiation and adherence among women in NYC and Philadelphia, we demonstrated feasibility and high acceptability of the Just4Us E&A intervention for increasing the uptake of daily oral PrEP among women. The Just4Us intervention is among the first theory-based interventions designed to help cisgender women make informed decisions about starting PrEP and the first to be evaluated in a randomized control trial[17].

Several other interventions have been evaluated among cisgender women in the US to increase PrEP uptake, indicating feasibility of various approaches, but with limited uptake In the pilot study evaluating a peer navigator-based intervention called PrEP-UP, which included cisgender and transgender women as well as transfeminine individuals, while 73% expressed interest in taking PrEP, and25% received help in scheduling PrEP appointments, none were prescribed PrEP[6]. In another study, a nurse-led PrEP program was associated with increased PrEP counseling compared to an active control using clinic decision support tools in an OB/GYN clinic; however, PrEP prescriptions were similar in both groups[9].

Our study findings of feasibility and high acceptability, along with the preliminary efficacy results previously described[17], are encouraging to move the Just4Us intervention into a future larger randomized trial to improve linkage to PrEP care and initiation of and adherence to PrEP among cisgender women in the US.

Our research team was successful in recruiting and enrolling cisgender women, with a large proportion identifying as Black and/or Latina, who experience higher burden of HIV infection, into the study. Of the 255 women recruited and assessed for eligibility for the study, 104 women (41%) were eligible, and 83 (80%) were enrolled in the two cities. We exceeded our target enrollment by race/ethnicity, with 26.3% of the women identifying as Hispanic/Latina and 78.8% identifying as Black/African American enrolled in the study. Our team employed a variety of strategies to recruit the women, including the successful referral process from enrolled women. Referrals from enrolled women incorporating a small monetary incentive could be used in the future to help recruit and enroll cisgender women who are at elevated risk for HIV infection.

The vast majority of those who were interested in PrEP but screened ineligible did not meet the criteria for PrEP eligibility according to the CDC screening guidelines in place at the time the study was conducted. However, the current CDC PrEP guidelines are more inclusive and allow individuals to self-determine if they are in need of PrEP without having to reveal to the provider which PrEP-relevant category they would meet[19]. This approach is consistent with reducing potentially stigmatizing impact of behavior-based screening to qualify for PrEP and has the potential to expand access to research and services involving biomedical HIV prevention[20].

The intervention completion rate in the study was high, with 100% of the modules completed. This is likely due to delivering the entire intervention content in one session. Satisfaction was also high among the women within the different components of the study. However, the length of the E&A session was noted by some women to be long. This could be adjusted by shortening some modules in a future trial, or by delivering the content over 2 or more sessions. However, this may result in lower levels of module completion.

Many follow-up phone call attempts were made, but calls were completed to only 39 participants, representing a completion rate of 65% (39/60) and indicating the study did not reach to the 90% benchmark for this component of the intervention. Among the calls that were completed, tailored adherence support was provided. Phone contact may be improved by having C-Ns call participants at non-traditional hours and outside of regular 9am to 5pm weekday schedules of the study sites. Access to mobile phones may be intermittent among socially disadvantaged populations, since almost 50% of the women in this RCT reported having phone service disconnected in the past years (at least once for some period of time) because she could not pay the bill or her phone was lost or stolen. Therefore, other outreach approaches are needed. These might include contacting friends or relatives or community locations where the women frequent, if such permission is provided ahead of time.

Enhanced navigation strategies are also likely needed. Exit interviews in our study[14] noted that some women reported substantial navigation barriers to PrEP care access (other than provider factors) (e.g., hard to schedule appointment around work, lost job and insurance, transportation issues, etc.) as well as concerns about seeing an unfamiliar provider. An intensified navigation component with a C-N will likely foster progression on the PrEP continuum among women. Enhanced navigation may include appointment scheduling, reminders, transportation assistance, accompanying women to an appointment (preferably with a PrEP competent provider), accompanying women to the pharmacy to fill PrEP prescription, actively assisting with accessing services to meet other needs such as jobs, health care insurance, childcare, etc. From our exit interviews, we learned that several women used the knowledge they gained from the Just4Us intervention to tell others in their community about PrEP. Such “PrEP ambassadors” may be a promising approach to increase community awareness of PrEP and could be incorporated into future intervention studies.

The automated text message program in the E&A arm was used by most participants, but there were limitations. Among those who received weekly messages, 85% described being satisfied/very satisfied with them. Among the 11% who started PrEP, only half indicated they received daily reminders. Thus, the text-messaging program was underutilized, which may have been due to the limited design and technical features. The text-message program was created because we were concerned some women in our sample might not have access to smartphones. However, smartphone access has increased over recent years even among our urban target population, with 98% of U.S. adults owning cell phones and 91% owning smartphones[21]; further, our baseline pilot RCT survey indicated 88% of participants did have a smartphone. In a future trial, instead of text-messaging, use of a professional-designed medication adherence smartphone app and customized to support PrEP navigation and adherence for women could be considered to provide adherence support. However, this may be challenging, as noted above, if women have limited or intermittent access to mobile phones.

At the time we developed the Just4Us intervention and conducted the pilot study, only one oral formulation was available for PrEP (i.e., TDF/FTC), but that has since changed. As noted above women expressed varied preference for PrEP modality, with 56% preferring a daily pill and 29% opting for injectable PrEP. The HPTN 084 trial demonstrated that cabotegravir (CAB), a long-acting injectable form of PrEP administered once every eight weeks, is safe and superior at preventing HIV acquisition among cisgender women in sub-Saharan Africa compared to the oral daily tenofovir disoproxil fumarate (TDF)/emtricitabine (FTC)[22]. In December 2021, the U.S. FDA approved injectable CAB for HIV prevention for adults and adolescents, including cisgender women[4]. Injectable CAB overcomes many of the barriers to medication adherence associated with daily TDF/FTC. Now that CAB LA for PrEP is an available option in the US, content about CAB LA should be incorporated into intervention strategies as appropriate and ensure PrEP provider referrals have the capacity to provide CAB LA to women. More long acting injectable PrEP options are on the horizon, as demonstrated by the recent Phase 3 PURPOSE 1 trial. The study showed 100% efficacy of subcutaneous lenacapavir, administered twice a year, in preventing HIV among adolescent girls and young cisgender women in Africa; this efficacy is higher than that of daily oral emtricitabine/tenofovir alafenamide (F/TAF) or daily oral TDF/FTC[23].

As noted in our previous paper[11], many women in our study chose to use condoms or abstain from sex as alternate ways to prevent HIV. In addition, with new biomedical HIV prevention now available, PrEP decision-making needs to be emphasized in future interventions for cisgender women and a more relevant endpoint for evaluation would be informed decision making, rather than PrEP uptake.

## Limitations

While the current study shows very promising results for the PrEP educational services, there is only retention information for 3 months following enrollment. Future research could include extending retention efforts and data for longer follow-up period of at least 6 months to track efficacy.

Women with limited literacy or non-English speakers are important include in PrEP health education programs, yet were not included in this study. In future research, PrEP and risk reduction content may be tailored specifically to populations that may be illiterate, have limited literacy, or may not be able to read and write in English, including translator services and/or interpreters.

The current analysis involves recruiting women mainly from homeless, shelters, through referrals and drug treatment centers and therefore cannot be generalized to other groups. Future research on the efficacy of the Just4Us educational intervention could focus on women at “lower risk”. This may include married women, middle and upper class women and other women that may have been missed during other outreach methods. For women considered “low risk”, there may be value to add discussions around HIV transmission in marriage or with a single, stable partner, regular check-ups, HIV screening despite perceived risk, and a review of the HIV modes of transmission.

Future studies may also include women under the age of 18. By including women outside of the specified age ranges in the inclusion-exclusion criteria, researchers may examine and revise the scope and content of the educational materials in the treatment group for a diverse range of audiences. The current educational materials could be revised and restructured based on different age cohorts that they’re designed to be administered to. For example, a social media component may be included for age cohorts under 18. In addition, our study specifically excluded women who are pregnant. Future studies should include pregnant women, given emerging evidence for safety of TDF/FTC as PrEP in pregnancy[24-26].

## Conclusions

The pilot study demonstrated feasibility and acceptability of the Just4Us E&A intervention, a promising woman-focused intervention to increase uptake of daily oral PrEP among cisgender women. These findings should be used to refine and test the adapted intervention in a future larger trial, especially with the availability of long-acting injectable PrEP.

## Declarations

Just4Us is funded by NIH/NIMH Intervention Development Grant (R34 #1R34MH108437-01A1; PI Anne Teitelman) and (P30AI045008; PI: Anne Teitelman).

We thank the Just4Us study team and the study participants who volunteered their time and energy to participate in this study.

## Clinicaltrials.gov registration

NCT03699722 A Women-Focused PrEP Intervention (Just4Us)

## Data Availability

All data produced in the present study are available upon reasonable request to the authors.

## References

1. Centers for Disease Control and Prevention. Estimated HIV incidence and prevalence in the United States, 2018–2022 in HIV Surveillance Supplemental Report 2024. 2024. https://www.cdc.gov/hiv-data/nhss/estimated-hiv-incidence-and-prevalence.html. Accessed 13 Sept 2024.

2. Raifman JR, et al. Brief Report: Pre-exposure Prophylaxis Awareness and Use Among Cisgender Women at a Sexually Transmitted Disease Clinic. J Acquir Immune Defic Syndr. 2019; 10.1097/qai.0000000000001879

3. U.S. Department of Health & Human Services. About Ending the HIV Epidemic in the U.S. 2023: https://www.hiv.gov/federal-response/ending-the-hiv-epidemic/overview/. Accessed 20 Jul 2023.

4. Food and Drug Administration. FDA News Release: FDA Approves First Injectable Treatment for HIV Pre-Exposure Prevention. 2021. https://www.fda.gov/news-events/press-announcements/fda-approves-first-injectable-treatment-hiv-pre-exposure-prevention. Accessed 3 May 2022.

5. Centers for Disease Control and Prevention. PrEP Coverage. 2023. https://www.cdc.gov/hiv/group/gender/women/prep-coverage.html. Accessed 4 Jan 2023.

6. Blackstock OJ, et al. A Pilot Study to Evaluate a Novel Pre-exposure Prophylaxis Peer Outreach and Navigation Intervention for Women at High Risk for HIV Infection. AIDS Behav. 2021; 10.1007/s10461-020-02979-y

7. O’Connell HR, Criniti SM. The Impact of HIV Pre-Exposure Prophylaxis (PrEP) Counseling on PrEP Knowledge and Attitudes Among Women Seeking Family Planning Care. J Womens Health (Larchmt). 2021; 10.1089/jwh.2019.8217

8. Roth AM, et al. Integrating HIV Preexposure Prophylaxis With Community-Based Syringe Services for Women Who Inject Drugs: Results From the Project SHE Demonstration Study. J Acquir Immune Defic Syndr. 2021; 10.1097/qai.0000000000002558

9. Wang R, et al. Clinic-based interventions to increase preexposure prophylaxis awareness and uptake among United States patients attending an obstetrics and gynecology clinic in Baltimore, Maryland. Am J Obstet Gynecol. 2023; 10.1016/j.ajog.2023.07.046

10. Teitelman A, Tieu HV, Davis A, Lipsky R, Darlington C, Iwu E, Brawne B, Shaw PA, Bond K, Frye V, Koblin BA. Just4Us: A Theory-Based PrEP Uptake Intervention Study for PrEP-Eligible Women in Two highly affected U.S. Cities with High Rates of HIV Shows High PrEP-Use Intentions but Many Barriers along the PrEP Cascade (2020, oral presentation), in *23rd International AIDS Conference (AIDS 2020: Virtual), July 6-10,* 2020.

11. Teitelman AM, Tieu HV, Chittamuru D, Shaw PA, Nandi V, Davis A, Lipsky R, Darlington C, Fiore D, Koblin BA. A Randomized Controlled Pilot Study of Just4Us, a Counseling and Navigation Intervention to Promote Oral HIV Prophylaxis Uptake Among PrEP-Eligible Cisgender Women. AIDS Behav. 2023; 10.1007/s10461-023-04017-z

12. Teitelman AM, et al. Just4Us: Development of a Counselor-Navigator and Text Message Intervention to Promote PrEP Uptake Among Cisgender Women at Elevated Risk for HIV. J Assoc Nurses AIDS Care. 2021; 10.1097/jnc.0000000000000233

13. Teitelman AM, et al. Individual, social and structural factors influencing PrEP uptake among cisgender women: a theory-informed elicitation study. AIDS Care. 2022; 10.1080/09540121.2021.1894319

14. Jackson GY, et al. Women’s views on communication with health care providers about pre-exposure prophylaxis (PrEP) for HIV prevention. Cult Health Sex. 2022; 10.1080/13691058.2021.1877824

15. Chittamuru D, et al. PrEP Stigma, HIV Stigma, and Intention to Use PrEP among Women in New York City and Philadelphia. Stigma Health. 2020; 10.1037/sah0000194

16. Ewing JA. Detecting alcoholism. The CAGE questionnaire. JAMA. 1984; 10.1001/jama.252.14.1905

17. Teitelman AM, et al. A Randomized Controlled Pilot Study of Just4Us, a Counseling and Navigation Intervention to Promote Oral HIV Prophylaxis Uptake Among PrEP-Eligible Cisgender Women. AIDS Behav. 2023; 10.1007/s10461-023-04017-z

18. Charmaz K, Constructing Grounded Theory. 2nd edition ed. Sage Publications; 2014.

19. Centers for Disease Control and Prevention. US Public Health Service: Preexposure prophylaxis for the prevention of HIV infection in the United States—2021 Update: a clinical practice guideline. 2021. https://www.cdc.gov/hiv/pdf/risk/prep/cdc-hiv-prep-guidelines-2021.pdf. Accessed 13 Sept 2024.

20. Golub SA, PrEP Stigma: Implicit and Explicit Drivers of Disparity. Curr HIV/AIDS Rep. 2018; 10.1007/s11904-018-0385-0

21. Pew Research Center. Mobile Fact Sheet. 2024. https://www.pewresearch.org/internet/fact-sheet/mobile/. Accessed 15 Feb 2024.

22. Delany-Moretlwe S, et al. Cabotegravir for the prevention of HIV-1 in women: results from HPTN 084, a phase 3, randomised clinical trial. Lancet. 2022; 10.1016/S0140-6736(22)00538-4

23. Bekker LG, et al. Twice-Yearly Lenacapavir or Daily F/TAF for HIV Prevention in Cisgender Women. N Engl J Med. 2024; 10.1056/nejmoa2407001

24. Joseph Davey D, Myer L, Coates T. PrEP implementation in pregnant and post-partum women. Lancet HIV. 2020; 10.1016/s2352-3018(19)30371-6

25. Joseph Davey D, et al. *W*here are the pregnant and breastfeeding women in new pre-exposure prophylaxis trials? The imperative to overcome the evidence gap. Lancet HIV. 2022; 10.1016/s2352-3018(21)00280-0

26. Bunge K, et al. DELIVER: A Safety Study of a Dapivirine Vaginal Ring and Oral PrEP for the Prevention of HIV During Pregnancy. J Acquir Immune Defic Syndr. 2024; 10.1097/qai.0000000000003312

